# Quantitative High-Frequency Ultrasound Identifies Spermatogenesis in Infertile Men with Non-Obstructive Azoospermia

**DOI:** 10.64898/2026.03.16.26348573

**Authors:** Taylor P. Kohn, Peyton J. Coady, Amelia G. Oppenheimer, Arnaav Walia, Beatriz S. Hernandez, Jaden R Kohn, Niki Parikh, Mahdi Bazzi, Blair T. Stocks, Mohit Khera, Larry I. Lipshultz

## Abstract

**Objective:** To determine whether quantitative ultrasound (QUS), which characterizes tissue microstructure using radiofrequency data, can identify regional heterogeneity within seminiferous tubules that corresponds to localized spermatogenesis in men with non-obstructive azoospermia (NOA).

**Design:** Two-cohort study using a biological extremes cohort to establish plausibility of a QUS biomarker, followed by an independent NOA-biopsy cohort with site-matched imaging and tissue sampling.

**Setting:** Academic male fertility referral center.

**Patients:** The biological extremes cohort included fertile men with presumed intact spermatogenesis (n=15) and men with NOA and subsequent negative microdissection testicular sperm extraction (mTESE; n=10). The NOA-biopsy cohort consisted of 27 men with NOA undergoing site-matched testicular biopsy via testicular sperm aspiration (TESA) or testicular sperm extraction (TESE), yielding 12 sperm-positive and 36 sperm-negative biopsy sites.

**Interventions:** High-frequency testicular ultrasound (36 MHz) with acquisition of raw radiofrequency data, allowing objective, quantitative analysis of tissue scattering patterns beyond conventional grayscale imaging. Regions of interest were manually annotated and, in the NOA-biopsy cohort, spatially matched to biopsy locations.

**Main Outcome Measures:** Association between sperm presence at biopsy sites and a pre-specified QUS measure of local tissue heterogeneity: the 75th percentile of a sliding window coefficient of variation map of the Nakagami k-factor within the superficial testicular parenchyma (K_Zone1_CV). This metric reflects the upper range of local variability in ultrasound backscatter, which is influenced by the underlying organization of seminiferous tubules.

**Results:** In the biological extremes cohort, K_Zone1_CV distinguished fertile controls (median 1.79, IQR 1.64-1.85) from NOA men with globally negative mTESE (median 1.51, IQR 1.42-1.58; P < 0.001), with an area under the receiver operating characteristic curve (AUC) of 0.91 (95% CI 0.79-1.00). In the independent NOA-biopsy cohort, K_Zone1_CV discriminated sperm-positive from sperm-negative biopsy sites with an AUC of 0.93 (95% CI 0.85-0.99). At a threshold of 1.60, sensitivity was 100%, specificity was 86.1%, positive predictive value was 70.6%, and negative predictive value was 100%. Serum hormone levels, testicular volumes, and biopsy technique did not differ significantly between groups.

**Conclusions:** Regional testicular tissue heterogeneity measured by quantitative ultrasound is associated with localized spermatogenesis in men with NOA. At the selected threshold, no sperm-positive biopsy site was misclassified as negative. These findings support the hypothesis that QUS can noninvasively detect the focal seminiferous tubule heterogeneity that predicts sperm retrieval success. This imaging approach could inform future image-guided sperm retrieval strategies. Further validation in larger cohorts and assessment of intra-patient variability are needed.

## INTRODUCTION

Non-obstructive azoospermia (NOA) affects one in a hundred men and represents 10-15% of cases in men seeking infertility care (1). After a diagnosis of NOA is made, the standard of care is to proceed with microdissection testicular sperm extraction (mTESE) where the testicle is bivalved and, under an operative microscope, the seminiferous tubules are systematically assessed, then the largest-diameter tubules are selectively sampled (2, 3). These larger-diameter tubules occur sporadically throughout the testicle in “islands” of preserved spermatogenesis, while the remainder consists of sclerotic tubules without sperm (3, 4). Successful sperm retrieval occurs in only 30-50% of patients, depending on the underlying pathology for NOA (e.g. Klinefelter syndrome, Y-chromosome microdeletion, etc.) (5). Despite advances in sperm retrieval techniques, there is no preoperative noninvasive method to quantify seminiferous tubule heterogeneity - the very characteristic that determines surgical success. Nor is there currently any imaging to guide a surgeon performing an mTESE on optimal locations to target sampling. This presents two clinical challenges or opportunities. First, if there are no islands of active spermatogenesis, could medical or hormonal optimization improve sperm production before proceeding to mTESE, leading to a higher chance of successful sperm retrieval? Second, akin to using seismic imaging technology to identify oil reserves prior to drilling - could high-frequency ultrasound and advanced data processing provide a map to the surgeon, ensuring that potential sperm-rich areas are targeted, ultimately leading to higher retrieval rates?

Recent studies have demonstrated that the key intraoperative predictor of successful sperm retrieval is the heterogeneity of seminiferous tubule distribution. Yu et al. found that men with heterogeneous tubule diameters under operative microscopy had a 65% sperm retrieval rate compared to 15% in those with homogeneous tubules (6). Similarly, Xiao et al. showed that testicular volume, as an indirect proxy for tubule distribution uniformity, was the only independent predictor of retrieval failure in idiopathic NOA (AUC 0.694), with larger, uniformly hyalinized testes predicting failure (7). However, both assessments require either surgery or prior biopsy, and no preoperative method exists to quantify this heterogeneity noninvasively.

Certainly, researchers have sought to assess whether various imaging modalities could identify spermatogenesis. Previous evaluation of CT or MRI to identify the presence of sperm have been attempted but have not been routinely used in clinical decision making (8). Recently, Nariyoshi et al. found that larger tubules could be identified through post-hoc manipulation of contrast and brightness of conventional ultrasound images; if at least one enlarged seminiferous tubule was found with this approach, there were significantly higher odds of finding sperm during mTESE (9). This preliminary exploration demonstrates that the enlarged tubules hunted during mTESE may be possible to identify using ultrasound. Advancing from this, it is reasonable to hypothesize that high-frequency ultrasound (HFUS) could provide greater differentiation of seminiferous tubules, as it has 3 times greater resolution. However, one prior study used HFUS to measure tubule size - evaluating NOA men with Sertoli cell only (SCO) versus men with obstructive azoospermia - and found no difference in tubule size when using high frequency ultrasound (10).

We hypothesized that additional, key tissue characteristics beyond tubule size may be detectable using quantitative ultrasound data. Conventional ultrasound produces traditional grayscale images, known as B-mode; these are qualitative in nature, are operator- and machine-dependent, and are liable to subjective visual interpretation. In contrast, quantitative ultrasound (QUS) analyzes the raw radiofrequency echoes to provide objective data about microstructural tissue properties. QUS has been used in several areas of medicine to differentiate tissue types that otherwise are imperceivable with traditional B-mode. QUS is particularly powerful in assessing tissue organization, cellular density, and spatial heterogeneity - which is uniquely suited to analyze testicular tissue. Seminiferous tubule histology is homogeneous in a patient with SCO (uniformly sclerotic), but heterogeneous in a patient with normal sperm (whose tubules are in various stages of spermatogenesis) (6, 7). This distinction between homogeneous and heterogeneous tissue architecture has been shown intraoperatively to predict mTESE success, but has never been quantified preoperatively.

The objective of this study was to determine whether local micro-heterogeneity within various testicular regions on quantitative ultrasound analysis is associated with the presence of sperm in men with non-obstructive azoospermia.

## METHODS

### Study Design

We retrospectively analyzed men presenting to an academic male fertility clinic between May 2024 and January 2026 under IRB approval. Using a previously published quantitative ultrasound (QUS) analysis feature found to correlate with total motile sperm count (11), we set out to test this QUS feature in two ways: first, comparing biological extremes – fertile men presenting for vasectomy and men who, after ultrasound, were found to have no sperm identified after mTESE; second, comparing NOA men with sperm-positive and sperm-negative biopsy sites in whom pre-operative ultrasound imaging was spatially matched to a single biopsy site. The primary analysis focused on a single pre-specified feature that assessed local spatial heterogeneity of ultrasound scattering properties rather than global testicular averages.

The first cohort was used to establish the biological plausibility of our QUS feature (11). This cohort included two control groups for comparison. First, we identified men with proven fertility within the past 3 years, who underwent an ultrasound just prior to undergoing vasectomy, and excluded any taking medications harmful to spermatogenesis (including testosterone therapy). Second, a group of men was used to classify ultrasound features associated with severe global spermatogenic failure. Specifically, we performed QUS on men who presented with NOA, and included in this group only those who had negative findings on subsequent mTESE (referred to below as “NOA-negative”).

The NOA biopsy cohort consisted of men with NOA undergoing testicular sperm extraction (TESE) or testicular sperm aspiration (TESA), in whom imaging could be spatially matched to a single biopsy site per patient. These NOA men presenting with infertility had serum hormone labs drawn and traditional scrotal ultrasounds performed at the time of presentation. To ensure men with obstructive azoospermia were not included, NOA was defined as FSH >7.6 IU/L, testicular length less <4.6 cm, and two semen analyses demonstrating azoospermia on pellet (12). In our academic clinical practice, all NOA men undergo an in-clinic TESA for predictive value prior to proceeding to mTESE (13). If an inadequate amount of tissue was obtained at the time of TESA, we reflex to an in-office TESE to obtain adequate tissue. Tissue is minced and then examined in our on-site andrology lab by both surgeon and andrology laboratory technicians. Prior to in-office TESA or TESE, NOA men included in the study underwent quantitative HFUS - HFUS was not used in real-time to change sperm retrieval practices; standard TESA or TESE was performed. After biopsy results returned, all NOA men in the NOA-biopsy cohort were assigned to either the “sperm-positive biopsy site” group or the “sperm-negative biopsy site” group.

Men were excluded if they had a prior testicular procedure, including TESE or mTESE, as scar tissue could misclassify tissue heterogeneity by affecting quantitative ultrasound RF scattering.

### Ultrasound Acquisition

HFUS imaging was performed by trained operators using a VevoMD high-frequency ultrasound (FUJIFILM VisualSonics, Toronto, Canada) with a 36-MHz linear array transducer with fixed acquisition parameters across all subjects. Raw ultrasound data were acquired in the in-phase and quadrature (IQ) format, allowing direct access to the underlying backscattered signal data, rather than relying on vendor-processed images. Imaging depth (∼14 mm), lateral field of view (∼15.36 mm), gain, and system presets were held constant to minimize technical variability between subjects.

### Rationale for Analytical Approach

This analysis was designed to reflect the biological reality that spermatogenesis is spatially heterogeneous. In men with infertility or sub-fertility, who may be able to produce sperm in focal areas, whole-testis averaging may obscure clinically relevant variation. By focusing on biopsy-matched superficial parenchyma and quantifying within-region variability, this approach aims to capture biologically meaningful differences associated with focal sperm production in men with NOA. This approach is motivated by intraoperative evidence that the spatial heterogeneity of seminiferous tubule diameters is a strong predictor of sperm retrieval success (6, 7).

### Quantitative Ultrasound Feature Extraction

QUS data enables the potential analysis of as many as 200 - 300 distinct features. For this study, our primary QUS feature was pre-specified based on prior exploratory analyses examining associations between QUS features and semen analysis parameters in an independent cohort (11). The prior exploratory analyses were used solely to guide feature selection and were not used for model training or validation in the present study. There was no overlap in the population associating QUS features with semen analyses and this cohort.

### Region of Interest

Regions of interest (ROIs) were manually annotated by a single investigator blinded to outcomes for both cohorts at the time of analysis. In the biological extremes cohort, multiple images (3-4 per testicle) were analyzed, and the ROI was drawn to reflect the entirety of the testicle with a depth of 3-4 mm and a total width of the captured testicle. In the NOA biopsy cohort, a single-site matched ultrasound image was captured and ROI were drawn with a depth of 3-4 mm and width of 3 mm to reflect the area biopsied. ROIs explicitly excluded the tunica albuginea, mediastinum testis, large vessels, and regions of acoustic shadowing on B-mode. ROIs were defined using Python package PyQtGraph and exported for quantitative analysis.

Within each ROI, a superficial parenchymal zone (Zone 1) was defined as the first 3-4 mm of testicular parenchyma deep to the tunica albuginea, excluding the capsule itself. This region was selected because (1) it corresponds to the typical depth sampled during TESE/TESA, (2) ultrasound resolution is highest near the surface, and (3) spermatogenesis is most likely to be preserved superficially in NOA (14). All primary analyses were restricted to this zone.

### Ultrasound Radiofrequency Processing

The ultrasound envelope measures the absolute value of the magnitude of the radiofrequency signal at each point in tissue – the envelope reflects the acoustic scattering properties at each pixel. B-mode images are a mathematically-transformed, grayscale, visual representation of the radiofrequency envelope data.

Ultrasound envelope data was reconstructed from the raw IQ (in-phase and quadrature) data using standard magnitude detection. To reduce variability related to coupling, attenuation, and gain, the envelope signal within each ROI was normalized by its median value. This normalization preserves relative spatial variation while minimizing non-biological differences between subjects (such as probe-skin coupling, beam incidence angle, near-field and focusing differences, or time-gain compensation).

The feature selected for analysis was derived from a measurement of local ultrasound scattering known as the Nakagami k-factor - a dimensionless parameter that describes how the envelope amplitudes are distributed (15). If the tissue is very heterogeneous, then the envelope amplitudes are quite variable within a single ROI (low k, with a high co-efficient of variation), whereas if the tissue is very homogeneous, then the amplitudes are similar or uniform within a single ROI (high k, with a low co-efficient of variation) (16, 17). Thus, the Nakagami k-factor reflects the spatial organization of a tissue and the variability of acoustic scattering within tissue (18). k-factor maps were computed using a local moment-based estimator with fixed kernel dimensions across all subjects.

After computing a k-factor map within zone 1, a sliding window (40 × 40 pixels) was passed across the zone, computing the local coefficient of variation (CV = σ/μ) of k-factor values at each position. A sliding window of 40 × 40 pixels (approximately 0.55 mm axial × 2.4 mm lateral) was used, spanning approximately 3–12 seminiferous tubule diameters per dimension. This produces a spatial map of local tissue heterogeneity, where each pixel represents how variable the scattering properties are in its immediate neighborhood. To summarize this map into a single biomarker per ROI, the 75th percentile of the local CV map captures local patches of heterogeneity that global measures dilute. The primary biomarker, K_Zone1_CV, was defined as the 75th percentile of this local CV map within zone 1.

In a prior exploratory study, we identified the coefficient of variation of the Nakagami k-factor within the superficial testicular parenchyma (K_Zone1_CV) as the QUS feature most strongly associated with sperm production (11). That analysis used a global summary statistic - pooling all k-factor values within zone 1 to compute a single CV. In the present study, we refined this approach by computing a sliding window CV map that captures local heterogeneity at each position within zone 1, and summarizing this map using the 75th percentile (SW-P75). This refinement was motivated by the recognition that focal spermatogenesis produces patchy regions of varied tubule architecture rather than uniformly elevated heterogeneity, and that a global average may dilute these focal signals (Supplemental Table 2).

### Statistical Analysis

In the biological extremes cohort, measurements from multiple images were aggregated at the individual patient level, using the median after analyzing 2-3 images per testicle in multiple ROIs. Differences in the coefficient of variation of the k-factor distribution between fertile controls and NOA-negative controls were assessed using the area under the receiver operating characteristic curve (AUC) and Precision-Recall Curves were used as the primary performance metric to evaluate K_Zone1_Cv and thresholds.

In the NOA-biopsy cohort, each patient contributed a one to two biopsy-matched ROI, and each biopsy site served as the unit of analysis. Validation performance metrics included AUC, Precision recall curves, sensitivity (If sperm exist, how often do we predict it), and specificity (If no sperm, how often do we predict it).

Uncertainty in AUC and average precision estimates was quantified using nonparametric bootstrap resampling (2,000 iterations; 95% confidence intervals; random seed = 42). Because patients in the NOA-biopsy cohort contributed a variable number of biopsy zones (range: 1–2 per patient), confidence intervals for that cohort were derived using a patient-level clustered bootstrap, in which patients, rather than individual zones, were resampled with replacement to preserve within-patient correlation structure (intraclass correlation coefficient = 0.54). Confidence intervals for the biological extremes cohort, in which each patient contributed a single summary value, were derived using standard nonparametric bootstrap resampling. Statistical significance was evaluated relative to a null hypothesis of AUC = 0.5.. Quantitative ultrasound image processing and statistical analyses were performed in Python; full details are provided in the Supplemental Methods.

## RESULTS

### Study Cohorts and Clinical Characteristics

A total of 52 men were included across biological extremes and NOA biopsy cohorts (Figure 1). The biological extremes cohort consisted of 15 fertile controls (intact spermatogenesis) and 10 men with non-obstructive azoospermia who had a negative mTESE (“NOA-negative,” those with severe global spermatogenic failure). The NOA biopsy cohort included 27 men, of whom 9 men had sperm identified on biopsy (for a total of 12 sperm-positive biopsies) and 18 men who had no sperm identified (36 sperm-negative biopsies). Clinical, endocrine, and testicular characteristics of NOA-negative men are found in Supplemental Table 1. Clinical, endocrine, and testicular characteristics of men in the NOA biopsy cohort are summarized in Table 1. Median serum follicle-stimulating hormone (FSH), luteinizing hormone (LH), total testosterone, and testicular volumes did not differ significantly between sperm-positive and sperm-negative biopsy groups. The proportion of patients undergoing each sperm retrieval technique was similar between groups (TESA vs TESE; Fisher’s exact test, p = 1.00), indicating that the biopsy method was unlikely to account for differences in sperm identification or imaging findings.

**Figure 1.**
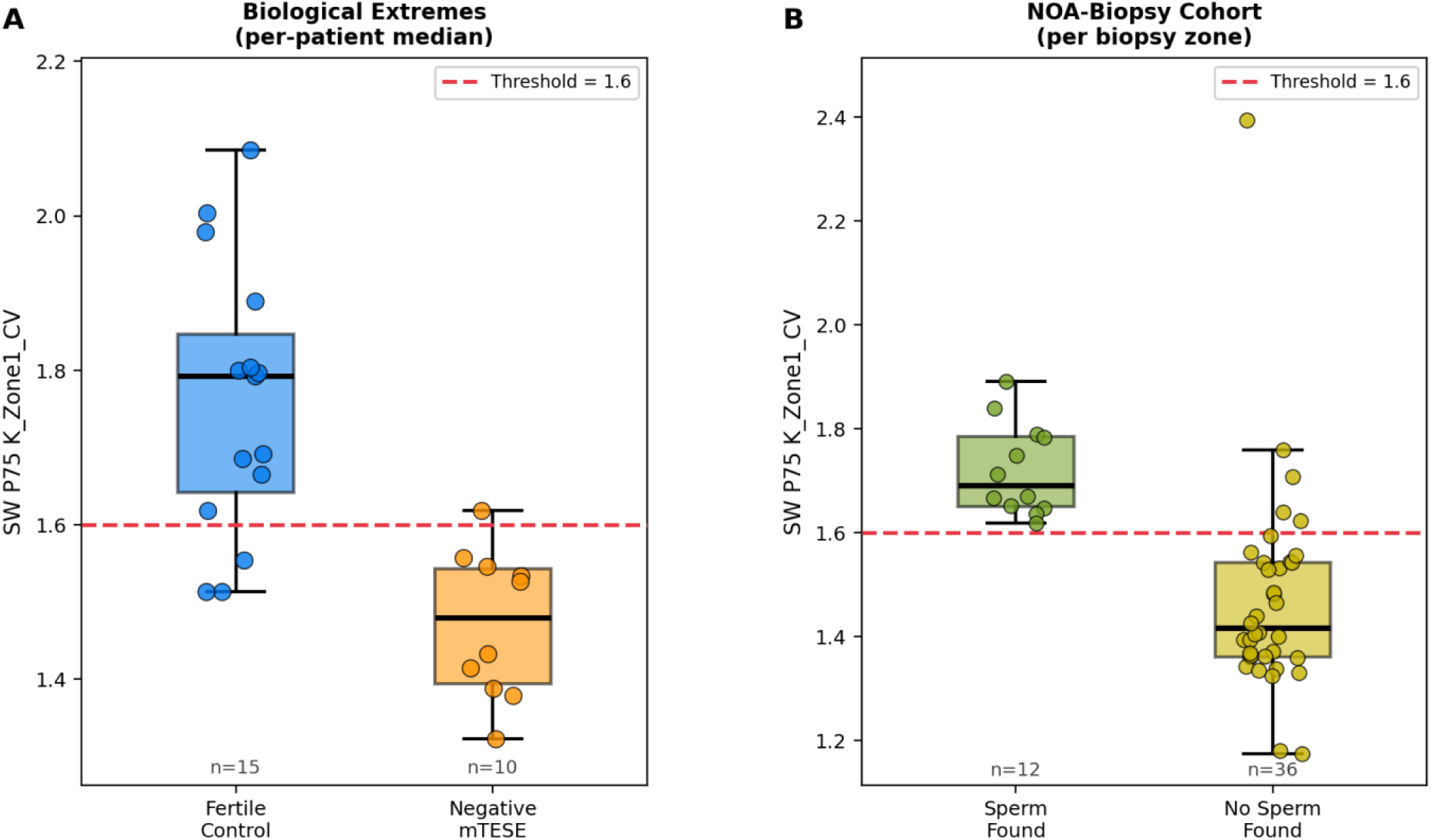
Boxplot Comparing Biological Extremes and NOA-Biopsy Cohorts: K_Zone1_CV discriminates spermatogenic status across two independent cohorts. (A) Per-patient median SW P75 K_Zone1_CV values in the Biological Extremes cohort (fertile controls, n=15; negative mTESE, n=10). Each point represents the median value across all biopsy zones for a given patient. (B) Per-zone SW P75 K_Zone1_CV values in the NOA-Biopsy cohort (sperm found, n=12 zones; no sperm found, n=36 zones). Each point represents a single biopsy zone. Boxes indicate the interquartile range (IQR) with the median shown as a horizontal line; whiskers extend to 1.5× IQR. The dashed red line denotes the unified diagnostic threshold of 1.6.

**Table 1.** Clinical and Endocrine Characteristics of Men in the NOA-Biopsy Cohort: Clinical, endocrine, and testicular characteristics of men with non-obstructive azoospermia (NOA) undergoing testicular biopsy. Values are reported as median with interquartile range (Q1–Q3). Testicular volumes were measured by ultrasound. Sperm retrieval technique was similar between sperm-positive and sperm-negative biopsy groups (Fisher’s exact test, p = 1.00).

| Variable | NOA undergoing Biopsy | NOA - Sperm Positive Biopsy | NOA - Sperm Negative Biopsy | P value |
| --- | --- | --- | --- | --- |
| **Number of Men** | **27** | **7** | **21** | **---** |
| **Biopsy Sites** | **48** | **12** | **36** |  |
| **FSH (IU/L)** | 15.9 (9.4–23.0) | 14.2 (10.5–21.9) | 16.2 (8.7–23.0) | 0.44 |
| **LH (IU/L)** | 10.3 (6.8–13.0) | 9.1 (7.5–12.4) | 10.7 (5.8–13.0) | 0.85 |
| **Total Testosterone (ng/dL)** | 349 (267–433) | 283 (250–379) | 373 (279–441) | 0.91 |
| **Left testicular volume (mL)** | 12.0 (6.9–14.0) | 12.7 (7.0–17.0) | 10.0 (5.9–13.5) | 0.21 |
| **Right testicular volume (mL)** | 12.0 (5.5–14.5) | 13.0 (9.0–16.0) | 9.0 (4.7–14.5) | 0.79 |
| **Sperm retrieval technique** | TESA: 39 (81%) TESE: 9 (19%) | TESA: 10 (83%) TESE: 2 (17%) | TESA: 29 (81%) TESE: 7 (19%) | 1.00 |

*median (quartile 1 - quartile 3)*

### Diagnostic Performance in the Biological Extremes Cohort

In the biological extremes cohort, K_Zone1_Cv was evaluated to assess whether global or regional testicular microstructural differences were detectable between fertile controls and men with NOA and negative mTESE. K_Zone1_Cv significantly discriminated fertile controls from NOA-negative men with fertile controls having a median of 1.79 (interquartile range 1.64 - 1.85) compared with NOA-negative men demonstrating a median of 1.51 (IQR: 1.42 - 1.58; p = 0.0006) (Figure 1A). AUC 0.91 (95% CI: 0.79-1.00; Figure 2A). Average precision on precision recall curve was 0.95 (95% CI: 0.85-1.00; Figure 2B). At the threshold of 1.6, sensitivity for identifying sperm-positive sites was 80%, with a specificity of 90% for identifying lack of sperm, PPV was 92%, and NPV was 75%.

**Figure 2.**
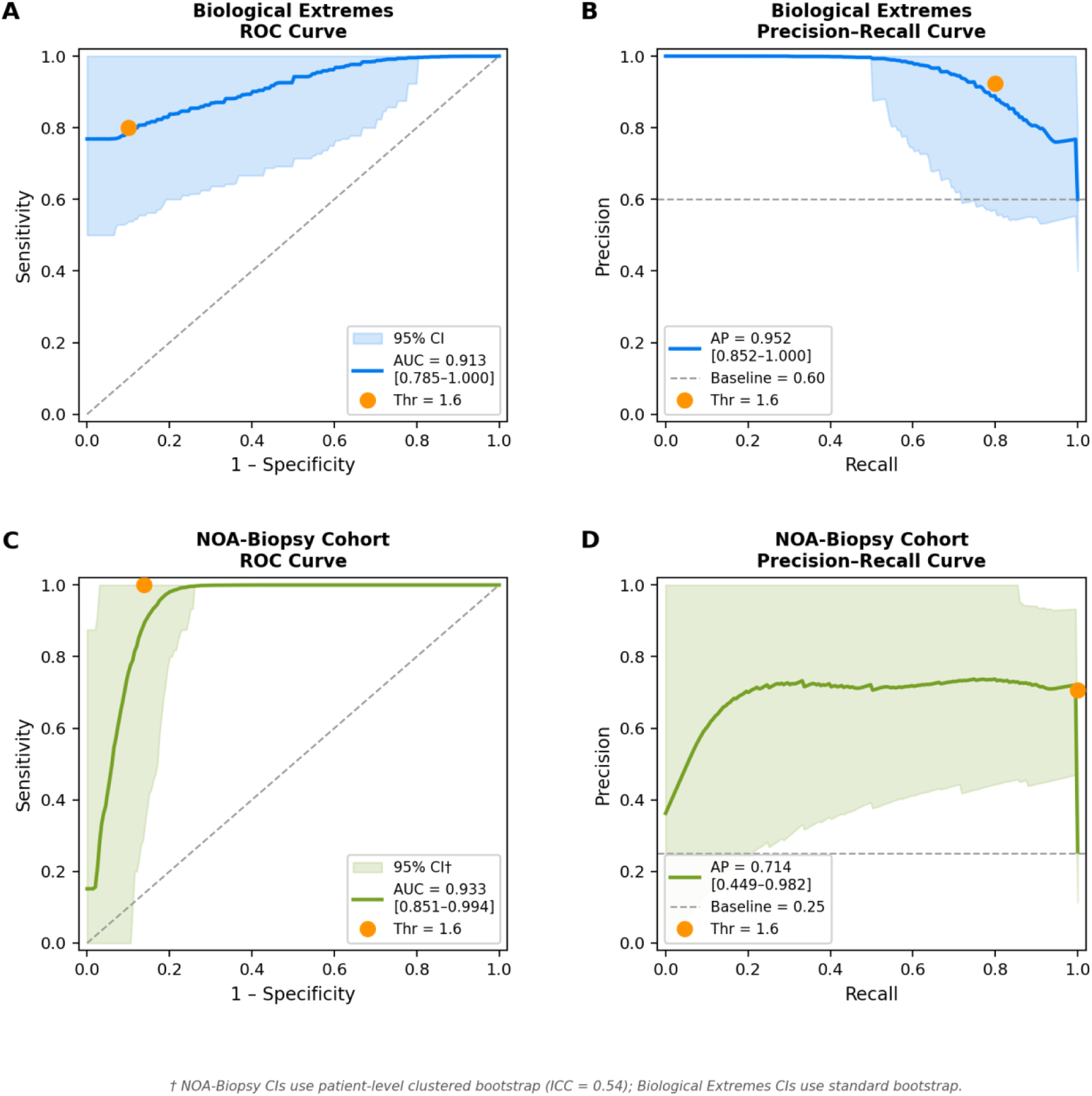
Diagnostic Performance of K_Zone1_Cv: Diagnostic performance of SW P75 K_Zone1_CV for predicting spermatogenic status. Receiver operating characteristic (ROC) and precision-recall (PR) curves for SW P75 K_Zone1_CV in the Biological Extremes cohort (A, B) and the NOA-Biopsy cohort (C, D). In the Biological Extremes cohort, classification was performed on per-patient median values (fertile controls vs. negative mTESE; n=25). In the NOA-Biopsy cohort, classification was performed at the per-zone level (sperm found vs. no sperm found; n=48 zones). Shaded regions represent 95% confidence intervals derived from 2,000 bootstrap iterations. The orange point marks performance at the pre-specified unified threshold of SW P75 ≥ 1.6. Dashed diagonal lines in ROC panels indicate chance-level performance; dashed horizontal lines in PR panels indicate the no-skill baseline. AUC, area under the ROC curve; AP, average precision.

### Site-Matched NOA Biopsy Cohort

K_Zone1_Cv was next evaluated in the NOA-biopsy cohort of men with NOA undergoing site-matched testicular biopsy to ensure spatial correspondence between imaging and histologic outcome. K_Zone1_Cv significantly discriminated NOA men with positive biopsies (median 1.69, IQR: 1.65-1.79) compared to NOA men with negative biopsies (median 1.42, IQR 1.36-1.54; p < 0.0001). In this cohort, K_Zone1_Cv discriminated sperm-positive from sperm-negative biopsy sites with an AUC of 0.93 (95% CI: 0.85–0.99; Figure 2C). Average precision was 0.71 (0.45 - 0.98). At the threshold of 1.6, sensitivity for identifying sperm-positive sites was 100%, with a specificity of 86% for identifying lack of sperm, PPV was 71% and NPV was 100% in the NOA-biopsy cohort.

Within-patient reliability of K_Zone1_CV across biopsy zones was quantified using a two-way mixed-effects ICC (absolute agreement, single measures); the estimated ICC of 0.54 (95% CI 0.18–0.79) indicated moderate within-patient correlation, supporting the use of a patient-level clustered bootstrap for confidence interval estimation in the NOA-biopsy cohort.

### Distribution of Superficial Zone Heterogeneity Across Clinical Groups

K_Zone1_Cv values across all four study groups demonstrated a clear gradient in superficial zone heterogeneity (Figure 3). Fertile controls exhibited the highest micro-structural heterogeneity, followed by sperm-positive NOA biopsy sites. In contrast, sperm-negative biopsy sites and NOA men with globally negative mTESE demonstrated lower and more homogeneous K_Zone1_Cv values – consistent with uniform, diffusely sclerotic tubules.

**Figure 3.**
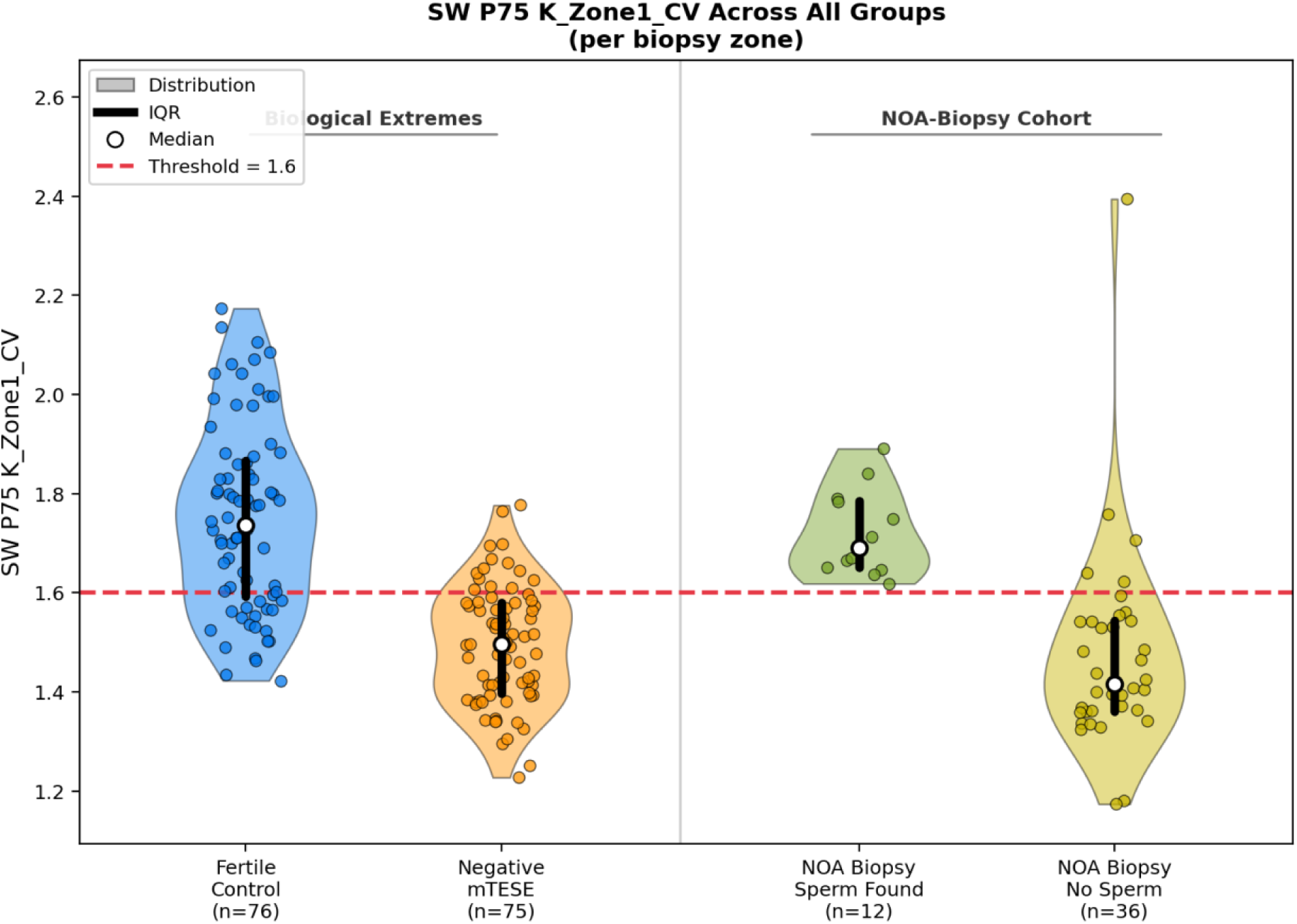
Distribution of Superficial Zone Heterogeneity Across Clinical Groups: Per-zone distribution of SW P75 K_Zone1_CV across all four groups. Violin plots showing the full distribution of SW P75 K_Zone1_CV values at the per-biopsy-zone level. The Biological Extremes cohort comprises fertile controls (n=76 zones, 15 patients) and negative mTESE patients (n=75 zones, 10 patients); the NOA-Biopsy cohort comprises biopsy sites where sperm were found (n=12 zones, 9 patients) and sites where no sperm were found (n=36 zones, 18 patients). Individual data points are overlaid with jitter to show data density. The central black bar indicates the interquartile range (IQR) and the white circle denotes the median. The dashed red line marks the unified diagnostic threshold of SW P75 K_Zone1_CV = 1.6.

Representative B-mode images and corresponding k-factor maps illustrated these differences qualitatively (Figure 4). Regions associated with sperm retrieval displayed visually heterogeneous patterns on the k-factor maps, whereas sperm-negative regions appeared more uniform, consistent with reduced microstructural complexity and diffusely sclerotic tubules.

**Figure 4.**
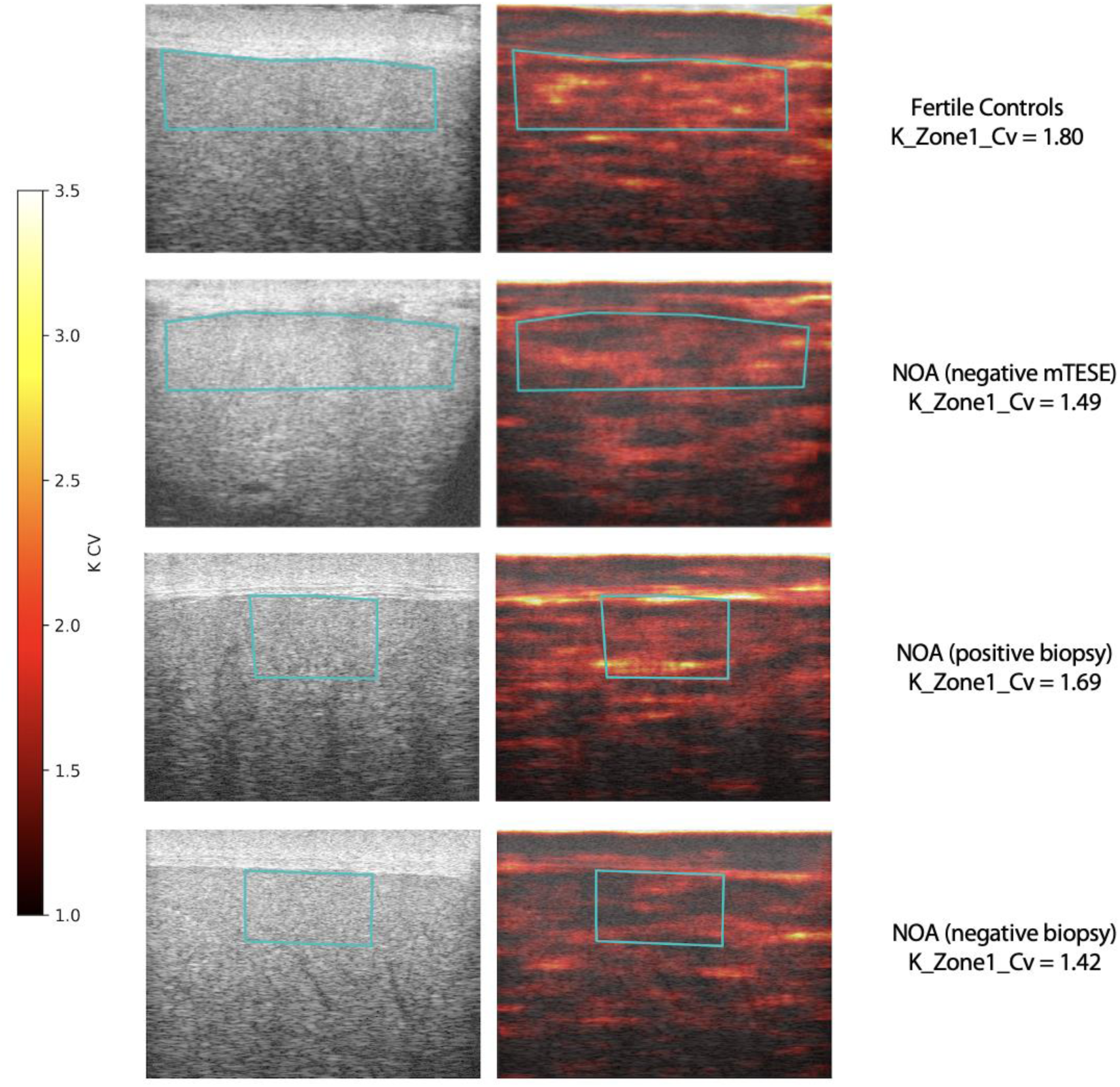
Representative Ultrasound Images and Heat Maps: Representative B-mode ultrasound images and corresponding k- Zone1-Cv maps. Regions of interest (ROIs) are specific for Zone 1 and are overlaid on B-mode images and corresponding maps. Fertile controls and sperm-positive biopsy sites demonstrate higher K_Zone1_Cv (higher heterogeneity), whereas sperm-negative biopsy sites and NOA men with negative mTESE exhibit lower K_Zone1_Cv (more homogeneous texture patterns).

## DISCUSSION

This is the first study that demonstrates quantitative ultrasound analyses can distinguish between the presence or absence of spermatogenesis-rich areas in NOA men, confirmed on biopsies. Importantly, this association was observed despite similar hormone profiles, testicular volumes, and biopsy techniques between groups, supporting the crucial importance of local tissue micro-architecture. This observation has a clear biological basis: healthy testes have complex seminiferous tubules with different cell types at various stages of sperm development. When sperm production fails, this complexity disappears and the tissue becomes more uniform and sclerotic, which is detectable as lower variation in heterogeneity on quantitative ultrasound measurements (6, 7). This is consistent with intraoperative observations by Yu et al. and Xiao et al., who demonstrated that heterogeneous seminiferous tubule distribution - assessed visually during mTESE or indirectly through testicular volume - was the strongest predictor of successful sperm retrieval (6, 7). Our findings suggest that K_Zone1_CV captures this same tissue characteristic noninvasively and preoperatively, with substantially higher discriminative accuracy than testicular volume alone (AUC 0.93 vs 0.69).

While the area under the ROC curve (AUC = 0.93) reflects strong overall discriminative ability, the precision-recall curve provides a more informative assessment of classifier performance in this setting given the inherent class imbalance - sperm-positive biopsy sites represented only 25% of all sites in the NOA-biopsy cohort. The average precision of 0.71 in the validation cohort, compared to 0.95 in the balanced biological extremes cohort, reflects this imbalance and underscores that positive predictive value will be sensitive to the prevalence of focal spermatogenesis in the population tested; future studies should report both metrics to allow meaningful cross-study comparisons.

While many studies have utilized various imaging modalities, ultrasound has several notable benefits: it can be cheaply performed, no exposure to radiation, easy transport, and can be used in real time to guide biopsies (19). Multiple high-frequency ultrasounds devices are FDA-approved, allowing for easy clinical implementation and could be used to guide routine patient care. With our current analysis strategy, imaging is not performed in real time; rather, mapping is done ahead of time, and then cognitive biopsies are performed. Planned future iterations will evaluate real-time mapping at the time of biopsy or sperm retrieval and will allow us to test intra-patient variability to better localize areas of spermatogenesis in a prospective fashion.

This study has several notable strengths. It was designed around a biologically-driven hypothesis that spermatogenesis in men with non-obstructive azoospermia is spatially heterogeneous, rather than uniformly distributed throughout the testis. Quantitative ultrasound analysis was performed using raw radiofrequency data acquired under standardized imaging conditions, and regions of interest were defined a priori to include only parenchyma and exclude the tunica, mediastinum, and vascular structures. In the NOA-biopsy cohort, imaging features were spatially matched to a single biopsy site per patient, ensuring close correspondence between imaging and clinical outcome as well as avoiding pseudo-replication. The primary feature evaluated has a clear biological interpretation and was able to discriminate between groups in the NOA-biopsy cohort, despite similar hormonal profiles, testicular volumes, and biopsy techniques between those groups.

Several limitations should be acknowledged. First, the sample size was modest, particularly in the sperm-positive NOA subgroup, which may limit statistical power for secondary analyses and precluded extensive multivariable modeling. However, effect sizes were consistent across biological extremes and NOA-biopsy cohorts, and performance estimates were supported by bootstrap confidence intervals. Second, fertile controls were assumed to be globally sperm-positive based on proven fertility rather than histologic confirmation. While this assumption is clinically reasonable, it introduces some uncertainty regarding microscopic heterogeneity in otherwise normal testes as fertile testes may still have micro-heterogeneity unrelated to spermatogenesis (6, 7). Comparing fertile men and men who had no sperm identified on mTESE represented biologic extremes. We hypothesized that if QUS could not distinguish between these two opposite groups, then it would not to be able to distinguish between areas of sperm or lack of sperm in NOA men.

Third, the study focused on superficial parenchymal regions to maintain consistent acoustic conditions and avoid attenuation artifacts. As a result, deeper testicular regions were not evaluated, and the findings may not fully capture heterogeneity at greater depths. Fourth, although biopsy technique (TESA versus TESE) was balanced between sperm-positive and sperm-negative groups, procedural factors such as needle trajectory or sampling volume were not explicitly modeled. Nevertheless, the absence of differences in technique distribution suggests that procedure type did not drive the observed associations. Comparison of NOA men was done in men undergoing biopsy rather than men undergoing mTESE since after bivalving we were not confident in the matching of the QUS locations identified preoperatively and intraoperative sampling. Finally, this study evaluated imaging features at the level of sperm presence or absence rather than detailed histopathologic subtypes of spermatogenic failure. Future prospective studies with larger cohorts and integrated, real-time histologic data, and sampling predicted positive and negative areas will be needed to solidify the clinical role of quantitative ultrasound in guiding sperm retrieval in men with NOA.

In this study, regional testicular microstructural heterogeneity measured by quantitative ultrasound was associated with localized spermatogenesis in men with non-obstructive azoospermia. By focusing on biopsy-matched superficial parenchyma, we identified a reproducible imaging feature that reflected local tissue variation and could discriminate between tissue with greater heterogeneity with likely active spermatogenesis versus the reduced heterogeneity of sclerotic tubules. These findings are consistent with a focal model of spermatogenesis and suggest that regional imaging analysis may provide valuable information in addition to conventional clinical assessment. While this approach may inform future image-guided sperm retrieval strategies and lays the groundwork for real-time mapping of spermatogenesis, it is not intended to replace intraoperative surgeon judgment during mTESE at this stage. Further studies are needed to validate these findings in a larger population and with multiple images and histologic samples collected per patient to assess intra-patient variability in men with non-obstructive azoospermia.

## Data Availability

All data produced in the present study are available upon reasonable request to the authors

---

**Supplemental Table 1:** Clinical and Endocrine Characteristics of Men With Non-Obstructive Azoospermia with subsequent Negative mTESE.

| Variable | NOA - No Sperm on mTESE |
| --- | --- |
| Number of Men | 10 |
| FSH (IU/L) | 13.4 (7.7–18.9) |
| LH (IU/L) | 8.9 (5.8–9.3) |
| Total Testosterone (ng/dL) | 362 (288–427) |
| Left testicular volume (mL) | 15.0 (10.3–17.0) |
| Right testicular volume (mL) | 13.5 (10.2–16.5) |

**Supplemental Table 2.** Clinical, endocrine, and testicular characteristics of men with non-obstructive azoospermia (NOA) who underwent mTESE and found to have no sperm. Values are reported as median with interquartile range (Q1–Q3). Testicular volumes were measured by ultrasound. Supplemental Table 2. Biomarker Summary Statistic Comparison and Threshold Selection **Supplemental Table 2A: Biomarker Summary Statistic Comparison: Biological Extremes Cohort** Five candidate summary statistics of the sliding window K-factor CV map were evaluated for their ability to distinguish fertile controls (n = 15 patients) from men with no sperm found on mTESE (n = 14 patients). Values represent patient-level medians. All five methods achieved 100% specificity (every mTESE-negative patient fell below threshold), reflecting the global nature of the difference between fertile and spermatogenically impaired tissue. AUC, sensitivity, specificity, and accuracy are reported at each method’s Youden-optimized threshold. Green shading indicates the summary statistic selected for validation in the NOA-biopsy cohort. **Supplemental Table 2B: Biomarker Summary Statistic Comparison: NOA-Biopsy Cohort** The same five candidate summary statistics were evaluated for their ability to distinguish sperm-positive from sperm-negative biopsy sites in men with NOA (12 positive sites in 10 patients; 36 negative sites in 17 patients). Each site represents a single spatially matched ultrasound-to-biopsy location. The sliding window 75th percentile (SW-P75) achieved the highest accuracy (89.6%) while maintaining 100% sensitivity — the clinically preferred operating point for a biomarker intended to avoid missing retrievable sperm. AUC, sensitivity, specificity, and accuracy are reported at each method’s Youden-optimized threshold. Green shading indicates the selected primary biomarker. **Supplemental Table 2C: Threshold Selection for SW-P75: NOA-Biopsy Cohort** Performance of the selected biomarker (SW-P75) across a range of classification thresholds in the NOA-biopsy cohort (12 sperm-positive sites, 36 sperm-negative sites). A threshold of ≥ 1.60 was selected as the operating point because it achieved the highest accuracy (89.6%) while preserving 100% sensitivity and 100% negative predictive value - ensuring that no sperm-positive site was misclassified as negative. The Youden index optimum (≥ 1.619, asterisk) achieved marginally higher specificity but at the cost of reduced sensitivity (91.7%). Green shading indicates the selected operating threshold.

| Method | Fertile Controls<br>Median [IQR] | mTESE Negative<br>Median [IQR] | AUC | Youden<br>Threshold | Sensitivity | Specificity | Accuracy |
| --- | --- | --- | --- | --- | --- | --- | --- |
| Global CV | 1.585 [1.468–1.663] | 1.354 [1.261–1.400] | 0.914 | 1.464 | 80.0% | 100.0% | 89.7% |
| SW Mean | 1.622 [1.495–1.691] | 1.375 [1.282–1.430] | 0.900 | 1.485 | 80.0% | 100.0% | 89.7% |
| SW Median | 1.576 [1.437–1.649] | 1.331 [1.268–1.396] | 0.900 | 1.459 | 66.7% | 100.0% | 82.8% |
| SW P75 | 1.793 [1.642–1.847] | 1.510 [1.419–1.579] | 0.900 | 1.665 | 73.3% | 100.0% | 86.2% |
| SW P95 | 2.211 [2.009–2.354] | 1.872 [1.659–1.913] | 0.914 | 1.977 | 80.0% | 100.0% | 89.7% |

| Method | Sperm Found<br>Median [IQR] | No Sperm<br>Median [IQR] | AUC | Youden<br>Threshold | Sensitivity | Specificity | Accuracy |
| --- | --- | --- | --- | --- | --- | --- | --- |
| Global CV | 1.491 [1.461–1.529] | 1.270 [1.213–1.355] | 0.921 | 1.382 | 100.0% | 83.3% | 87.5% |
| SW Mean | — | — | 0.942 | 1.410 | 100.0% | 80.6% | 85.4% |
| SW Median | — | — | 0.905 | 1.358 | 91.7% | 77.8% | 81.3% |
| SW P75 | 1.691 [1.651–1.785] | 1.417 [1.361–1.543] | 0.933 | 1.619 | 100.0% | 86.1% | 89.6% |
| SW P95 | — | — | 0.898 | 1.972 | 100.0% | 83.3% | 87.5% |

| Threshold | TP | FN | TN | FP | Sensitivity | Specificity | PPV | NPV | Accuracy |
| --- | --- | --- | --- | --- | --- | --- | --- | --- | --- |
| ≥ 1.50 | 12 | 0 | 23 | 13 | 100.0% | 63.9% | 48.0% | 100.0% | 72.9% |
| ≥ 1.55 | 12 | 0 | 28 | 8 | 100.0% | 77.8% | 60.0% | 100.0% | 83.3% |
| ≥ 1.58 | 12 | 0 | 30 | 6 | 100.0% | 83.3% | 66.7% | 100.0% | 87.5% |
| ≥ 1.60 | 12 | 0 | 31 | 5 | 100.0% | 86.1% | 70.6% | 100.0% | 89.6% |
| ≥ 1.619* | 11 | 1 | 31 | 5 | 91.7% | 86.1% | 68.8% | 96.9% | 87.5% |
| ≥ 1.63 | 11 | 1 | 32 | 4 | 91.7% | 88.9% | 73.3% | 97.0% | 89.6% |
| ≥ 1.65 | 9 | 3 | 33 | 3 | 75.0% | 91.7% | 75.0% | 91.7% | 87.5% |
| ≥ 1.70 | 6 | 6 | 33 | 3 | 50.0% | 91.7% | 66.7% | 84.6% | 81.2% |

Green row = selected operating threshold (≥ 1.60)
\* Youden's index optimum
Threshold ≥ 1.60 selected to maximize accuracy while preserving 100% sensitivity

## Supplemental Methods – Software

All quantitative ultrasound analyses were performed using custom-written scripts in Python (version 3.13.2; Python Software Foundation). Raw ultrasound in-phase and quadrature (IQ) data were processed to generate envelope images, from which quantitative ultrasound features were derived. Image processing, numerical computation, and statistical analyses were implemented using standard open-source scientific computing libraries, including NumPy, SciPy, scikit-image, and pandas. Data visualization was performed using matplotlib.

Regions of interest were manually annotated using PyQtGraph and exported as GeoJSON files for downstream analysis. Bootstrap resampling, receiver operating characteristic analyses, precision recall curves, and nonparametric statistical tests were implemented within the Python environment using reproducible random seeds. All analyses were conducted on de-identified data.

